# Nasal Epigenome-Wide Association Study of Respiratory and Allergic Biomarkers in Mexican American Young Adults

**DOI:** 10.64898/2026.10.27.26366640

**Authors:** Javier Perez-Garcia, Robert B. Gunier, Brenda Eskenazi, John Balmes, Katherine Kogut, Weihong Guo, Sheryl L Rifas-Shiman, Marie-France Hivert, Emily Oken, Diane R. Gold, Stephanie Holm, Nina Holland, Andres Cardenas

## Abstract

**Background:** Mexican Americans have low asthma prevalence, but they experience a disproportionate clinical asthma burden. Nasal DNA methylation (DNAm) is a biomarker of airway diseases, but its relevance in young adult Mexican Americans and links to air pollution are underexplored.

**Objective:** To identify nasal DNAm markers associated with asthma, allergic sensitization, fractional exhaled nitric oxide (FeNO), and lung function and evaluate associations with air pollution.

**Methods:** We tested associations between nasal DNAm at 804,220 CpGs (EPICv2) and respiratory outcomes in 354 young adults from CHAMACOS (age 22) and evaluated replication in 540 adolescents from Project Viva (age 13). Epigenome-wide association studies were adjusted for confounders, and significance was declared at a false discovery rate (FDR)<0.05. Significant CpGs were evaluated for association with prenatal and past-year PM_2.5_ and O_3_ exposures, and with gene expression and genetic variation (*in silico*). We examined differentially methylated regions (DMRs), gene-set enrichments, and epigenetic aging.

**Results:** We identified 49 CpGs associated with FeNO and 557 DMRs across multiple respiratory outcomes (FDR<0.05). Twenty-six CpGs and 103 DMRs associated with FeNO, asthma, and FEV_1_/FVC were replicated (e.g., *LRRFIP1*, *PCSK6*, *EPX, IL13*). Ten FeNO-related CpGs were associated with PM_2.5_ and/or O_3_ exposures (*p*<0.05) and 15 CpGs with nasal epithelial expression. We found enrichment for associations with respiratory/allergic biomarkers, obesity, smoking, longevity, asthma-related pathways (PI3K-AKT, TNF-α, IL-2), inflammatory transcription factors, and candidate asthma drugs. FeNO was associated with greater epigenetic aging acceleration (β=0.3, *p*=0.008).

**Conclusions:** We identified nasal DNAm markers implicated in airway epithelial integrity and immune response, predominantly associated with FeNO.

## INTRODUCTION

Asthma is a heterogeneous respiratory syndrome characterized by chronic airway inflammation and reversible airflow limitation, often accompanied by allergic sensitization, eosinophilia, lung function decline, increased inflammatory biomarkers, or high fractional exhaled nitric oxide (FeNO) (1). In the U.S., there are 27.8 million people with asthma (2), representing the most prevalent chronic pediatric disease. It contributes to a reduced quality of life for patients and families, and uncontrolled asthma will cost the U.S. economy an estimated $963 billion in the following two decades (3). Asthma disparities across different ethnic groups are well documented. Although lower asthma prevalence is reported among Mexican Americans compared to other populations, they suffer disproportionately high burden of uncontrolled asthma and exacerbations (4).

Asthma results from the interaction of genetic factors (heritability estimated at 40-70%) and multiple environmental factors (*e.g.,* air pollution, smoking, microbes, and allergens), whose exposures during pre- and postnatal life stages contribute to disease onset and severity (5). Based on this, the epigenome, which reflects both factors, is key in understanding asthma pathology and biomarker discovery (5–9). DNA methylation (DNAm), the methylation of cytosines often located in 5’-cytosine-phosphate-guanine-3’ dinucleotides (CpG sites), has been the epigenetic marker most studied in asthma over the past decade (5–9).

Multiple epigenome-wide association studies (EWAS) have discovered thousands of CpGs and differentially methylated regions (DMRs) associated with asthma and other allergies (8,7). Although most EWAS have focused on blood cells, several studies have analyzed nasal samples given that nasal mucosa is the first line of the airways in contact with the airborne environment, and the nasal epithelial barrier is key for protection and clearance of microbial pathogens, particles, and toxins (8,7). Nasal epithelial DNAm markers associated with asthma risk mainly participate in airway inflammation, immune system regulation, eosinophilic response, and epithelial barrier integrity (8,7). Furthermore, the nasal methylome can serve as a source of biomarkers of asthma, atopy, and respiratory health (6,10).

Most epigenomic studies of asthma have focused on children (8,7), and it remains unclear whether nasal DNAm biomarkers persist into adulthood. Moreover, although blood DNAm has been proposed as a mediator of the association between air pollution exposure and asthma control (11), the overlap of nasal DNAm signatures between asthma and air pollution has not been explored. Finally, the most comprehensive Illumina MethylationEPICv2 BeadChip to date, which interrogates ∼950,000 CpG sites, has not yet been applied in EWAS of asthma using nasal samples.

We hypothesize that the nasal epigenome is associated with respiratory health in Mexican American young adults. We analyzed 354 participants to identify CpGs and DMRs associated with asthma (current and ever diagnosis, and early and late onset), allergic sensitization, fractional exhaled nitric oxide [FeNO], and several spirometry measurements. Identified signals were followed up for replication in an independent multi-ethnic cohort of 540 adolescents and tested for association with air pollution exposure. We conducted *in silico* associations with gene expression and genetic variations, examined enrichment in biological pathways, traits, and drug signatures, and analyzed associations with epigenetic aging.

## METHODS

An extensive Methods section is reported in the **Supplemental Material**.

### Study Populations

In the discovery phase, we analyzed 354 young adults from the Center for the Health Assessment of Mothers and Children of Salinas (CHAMACOS) cohort study (12,13). CHAMACOS is a longitudinal birth cohort investigating the effects of prenatal and early postnatal environmental exposures on the health of Mexican American children and young adults living in California’s Salinas Valley. Pregnant mothers and their children were followed up at 1- to 2-year intervals, and 440 participants were followed up for the young adult visit (age 23).

In the replication phase, we analyzed 540 nasal samples from adolescents (age 13) from the pre-birth multiethnic cohort Project Viva. A full description of the cohort design, nasal epigenome-wide DNAm profiling, and respiratory outcomes assessment was described elsewhere (6,14,15).

Informed consent and assent were obtained from mothers and children, as appropriate. All protocols obtained ethical approvals (The University of California, Berkeley Committee for the Protection of Human Subjects for CHAMACOS, and the IRB of Harvard Pilgrim Health Care for Project Viva).

### Respiratory Health Assessment

Asthma was defined as a reported doctor’s diagnosis of asthma anytime from birth through the 22-year visit (reported either by the participant or their mother). We defined asthma by age of onset as early (≤9 years old) or late (>9 years old). We defined ongoing asthma as having both an early asthma diagnosis and asthma symptoms or medications in the past year. Lung function parameters, including forced expiratory volume in one second (FEV_1_), forced vital capacity (FVC), the forced expiratory flow from 25% to 75% of FVC (FEF_25-75_), peak expiratory flow (PEF), and FEV_1_/FVC, as well as FeNO, were measured by trained staff following established guidelines (16,17). Results were independently reviewed by two physicians, with discrepancies resolved collaboratively. Allergic sensitization was defined based on total IgE levels ≥200 IU/ml measured at age 22.

### Genome-wide DNAm measurement and quality control

Trained research assistants collected nasal swabs from the anterior nares of 438 young adults, of which 359 (82%) yielded adequate respiratory epithelial cells for epigenetic laboratory analysis. We measured DNAm at 936,990 CpGs across the genome using the Infinium Illumina MethylationEPICv2 BeadChip (Illumina, San Diego, CA). A standardized quality control (QC) of DNAm data was conducted using the *minfi*, *ENmix*, and *ewastools* R packages (18–20). Briefly, we corrected background noise and dye and probe-type biases, and inter-array variation. We filtered out low-quality probes and those with mapping inaccuracies, low-performance issues, missingness rate >5%, non-annotated to autosomes, replicated probes, and cross-reactive (21,22) and potentially polymorphic. We also filtered out two potentially cross-sample contaminated individuals. Beta-values were corrected for batch using ComBat-met and transformed to M-values to minimize skewed distributions in subsequent analyses (23,24).

### Statistical analyses

#### Epigenome-wide association study

We tested associations between DNAm levels at CpGs and respiratory outcomes through linear regression models adjusted for age, sex, race and ethnicity, body mass index (BMI) (z-score), household income category, smoking and/or vaping in the past month, season of sample collection (sine and cosine), and tissue heterogeneity (ReFACTor components) (25). EWAS were conducted independently for each respiratory outcome, including asthma (ever), current asthma, early-onset asthma, late-onset asthma, total IgE ≥200 IU/ml, FEV_1_ (z-score), FVC (z-score), FEV_1_/FVC, PEF (z-score), and FEF_25-75_ (z-score). DMRs were estimated using two independent methods: *ipDMR* and comb-p (18,26,27). We retained DMRs that contained≥2 CpGs and were significantly detected using both methods. Significance was declared at a false discovery rate (FDR)<0.05.

#### Replication of CpGs and DMRs in Project Viva

CpGs and DMRs significantly associated with any respiratory outcome in CHAMACOS (FDR<0.05) were followed up for replication in 540 nasal samples from adolescents from Project Viva. Each CpG or DMR was tested only for its association with the same respiratory outcome identified in the discovery analysis. Replication was declared at *p*<0.05.

#### DNAm associations with air pollution exposure

As described in the **Supplementary Methods**, based on EPA monitoring stations and geocoded residential addresses, we estimated time-weighted PM_2.5_ (µg/m^3^) and O_3_ (ppb) average prenatal and past-year exposures. We tested in CHAMACOS associations between DNAm at selected CpGs associated with respiratory outcomes and log_2_-transformed air pollution exposure using linear regression models adjusting for the same covariates as in the EWAS.

#### *In silico* quantitative trait loci analyses

We inspected whether identified CpGs had been associated with nasal epithelia gene expression (expression quantitative trait methylation, eQTM) and/or with genetic variation (methylation quantitative trait loci, meQTL) using a public database from 455 Puerto Rican youth with and without atopic asthma (28).

#### Enrichment analyses

We conducted gene-set enrichment analyses to identify enrichment in biological pathways, gene ontologies, human diseases/traits, transcription factor binding sites, and/or drug signatures using Enrichr (29). We analyzed gene annotations for suggestive signals (FDR<0.2) from any respiratory outcome in the discovery phase. To ensure robustness of associations, we re-assessed the analysis using different arbitrary FDR cutoffs (FDR<0.3 and FDR<0.1).

#### Epigenetic aging

We tested associations between the Horvath and Skin&Blood epigenetic clocks (DNAm biomarkers predictive of chronological age) and respiratory outcomes using linear regression models adjusted for the same covariates as in the EWAS. Significance was declared for each outcome at *p*<0.025 (α=0.05/2 epigenetic clocks=0.025).

## RESULTS

### Study population

We analyzed 354 young adults with available nasal epigenomic data from CHAMACOS in the discovery phase. The main demographic and clinical characteristics of the study sample are reported in **Table 1**. Briefly, young adults had a mean (SD) age of 22.5 (0.7) years, and 57.3% were female. Most self-identified as Mexicans (89.0%), with smaller proportions of other Latinos (8.5%) or other race/ethnicity (2.5%). Most participants had obesity (43.2%) or were overweight (29.4%), 72.9% had a household income below the poverty line, and 35.3% smoked or vaped in the past month. A subset of 99 (28.0%) participants had ever had asthma, 55 (15.5%) had a current asthma diagnosis, and 118 (34.8%) had total IgE levels >200 UI/ml. For respiratory/allergic biomarkers, the mean (SD) for FeNO was 20.1 (21.3) ppb, for FEV_1_ (z-score) was −0.3 (1.0), for FVC (z-score) was −0.04 (1.1), and for FEV_1_/FVC was 0.9 (0.1).

**Table 1.**
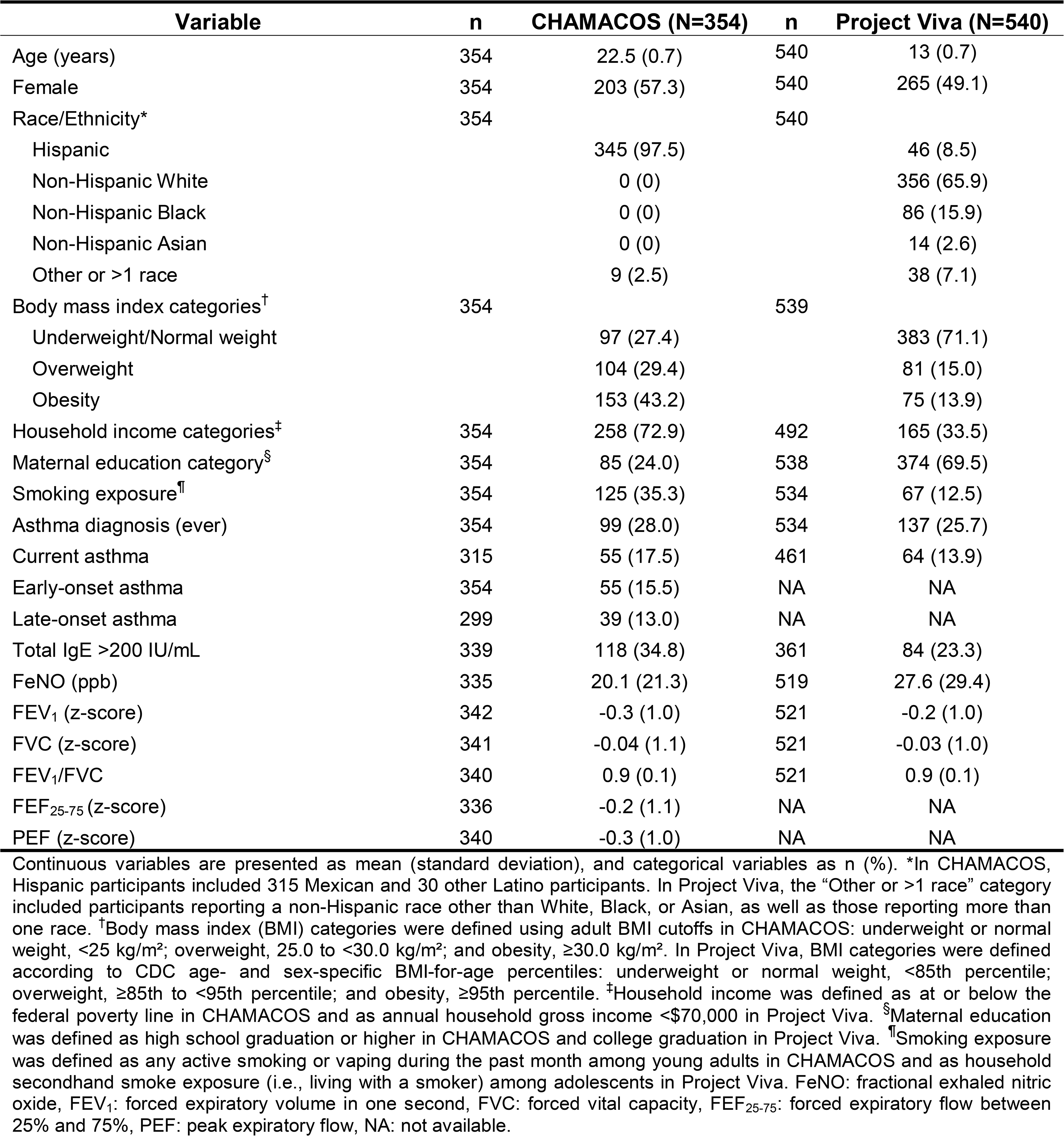
Demographic and clinical characteristics of the study participants.

| Variable | n | CHAMACOS (N=354) | n | Project Viva (N=540) |
| --- | --- | --- | --- | --- |
| Age (years) | 354 | 22.5 (0.7) | 540 | 13 (0.7) |
| Female | 354 | 203 (57.3) | 540 | 265 (49.1) |
| Race/Ethnicity* | 354 |  | 540 |  |
| Hispanic |  | 345 (97.5) |  | 46 (8.5) |
| Non-Hispanic White |  | 0 (0) |  | 356 (65.9) |
| Non-Hispanic Black |  | 0 (0) |  | 86 (15.9) |
| Non-Hispanic Asian |  | 0 (0) |  | 14 (2.6) |
| Other or >1 race |  | 9 (2.5) |  | 38 (7.1) |
| Body mass index categories<sup>†</sup> | 354 |  | 539 |  |
| Underweight/Normal weight |  | 97 (27.4) |  | 383 (71.1) |
| Overweight |  | 104 (29.4) |  | 81 (15.0) |
| Obesity |  | 153 (43.2) |  | 75 (13.9) |
| Household income categories<sup>‡</sup> | 354 | 258 (72.9) | 492 | 165 (33.5) |
| Maternal education category<sup>§</sup> | 354 | 85 (24.0) | 538 | 374 (69.5) |
| Smoking exposure<sup>¶</sup> | 354 | 125 (35.3) | 534 | 67 (12.5) |
| Asthma diagnosis (ever) | 354 | 99 (28.0) | 534 | 137 (25.7) |
| Current asthma | 315 | 55 (17.5) | 461 | 64 (13.9) |
| Early-onset asthma | 354 | 55 (15.5) | NA | NA |
| Late-onset asthma | 299 | 39 (13.0) | NA | NA |
| Total IgE >200 IU/mL | 339 | 118 (34.8) | 361 | 84 (23.3) |
| FeNO (ppb) | 335 | 20.1 (21.3) | 519 | 27.6 (29.4) |
| FEV<sub>1</sub> (z-score) | 342 | -0.3 (1.0) | 521 | -0.2 (1.0) |
| FVC (z-score) | 341 | -0.04 (1.1) | 521 | -0.03 (1.0) |
| FEV<sub>1</sub>/FVC | 340 | 0.9 (0.1) | 521 | 0.9 (0.1) |
| FEF<sub>25-75</sub> (z-score) | 336 | -0.2 (1.1) | NA | NA |
| PEF (z-score) | 340 | -0.3 (1.0) | NA | NA |

### EWAS of respiratory outcomes

Among all respiratory outcomes analyzed (**Table 2, Figure S1**), we identified 49 CpGs significantly associated with FeNO levels (λ=1.18, FDR<0.05, **Figure 1**, **Table 3**). The most significant association was with the CpG cg02797113 annotated to *LRRFIP1*, whose DNAm levels were associated with lower FeNO (logFC = −0.20, *p =* 1.2×10^−8^). Among the 49 CpG sites, annotated genes were involved in epithelial integrity (*TFF3*, *EHF*, *SLC9A3*), eosinophil recruitment (*CCL26*), immune system regulation (*CD200R1*, *BATF3*), and allergic inflammation (*NTRK1*, *PCSK6*). We did not detect any other associations between nasal DNAm at individual CpGs and other respiratory outcomes.

**Figure 1.**
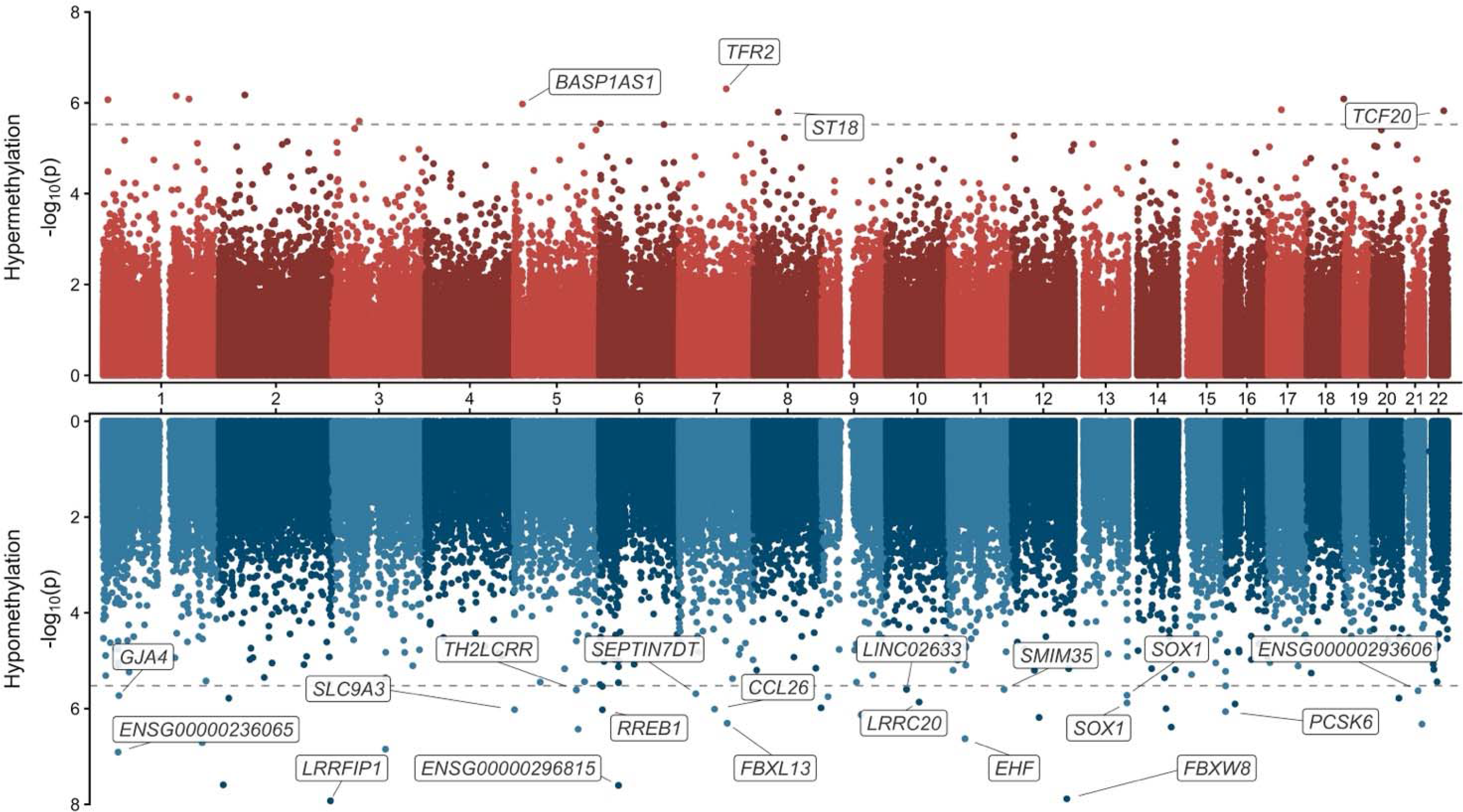
Miami plot of the EWAS of FeNO in the discovery phase (CHAMACOS). Association -log_10_(*p*) is represented on the y-axis and genomic coordinates on the x-axis. CpGs hypermethylated in cases are represented in the upper panel (red) and those hypermethylated in the lower panel (blue). The gray dashed line represents the genome-wide significance threshold (*p*<9×10^−8^). Gene annotation is provided for CpGs with evidence of replication in Project Viva.

**Table 2.** Summary nasal EWAS of respiratory outcomes.

| **Outcome** | **N** | **$\lambda_{GC}^*$** | **CpGs (FDR<0.05)** | **DMRs (FDR<0.05)** |
| --- | --- | --- | --- | --- |
| Asthma (ever) | 354 | 1.00 | 0 | 1 |
| Current asthma | 315 | 1.05 | 0 | 3 |
| Early-onset asthma | 354 | 1.06 | 0 | 5 |
| Late-onset asthma | 299 | 0.92 | 0 | 4 |
| Allergic sensitization (IgE>200 IU/ml) | 339 | 1.05 | 0 | 6 |
| FeNO | 335 | 1.18 | 49 | 327 |
| FEV<sub>1</sub> (z-score) | 342 | 1.02 | 0 | 8 |
| FVC (z-score) | 341 | 0.97 | 0 | 4 |
| FEV<sub>1</sub>/FVC | 340 | 1.15 | 0 | 86 |
| FEF<sub>25-75</sub> (z-score) | 336 | 1.15 | 0 | 111 |
| PEF (z-score) | 342 | 1.01 | 0 | 2 |

\*Genomic inflation. FDR: false discovery rate, IgE: Immunoglobulin E, FeNO: fractional exhaled nitric oxide, FEV<sub>1</sub>: forced expiratory volume in one second, FVC: forced vital capacity, FEF<sub>25-75</sub>: forced expiratory flow between 25% and 75%, PEF: peak expiratory flow.

**Table 3.**
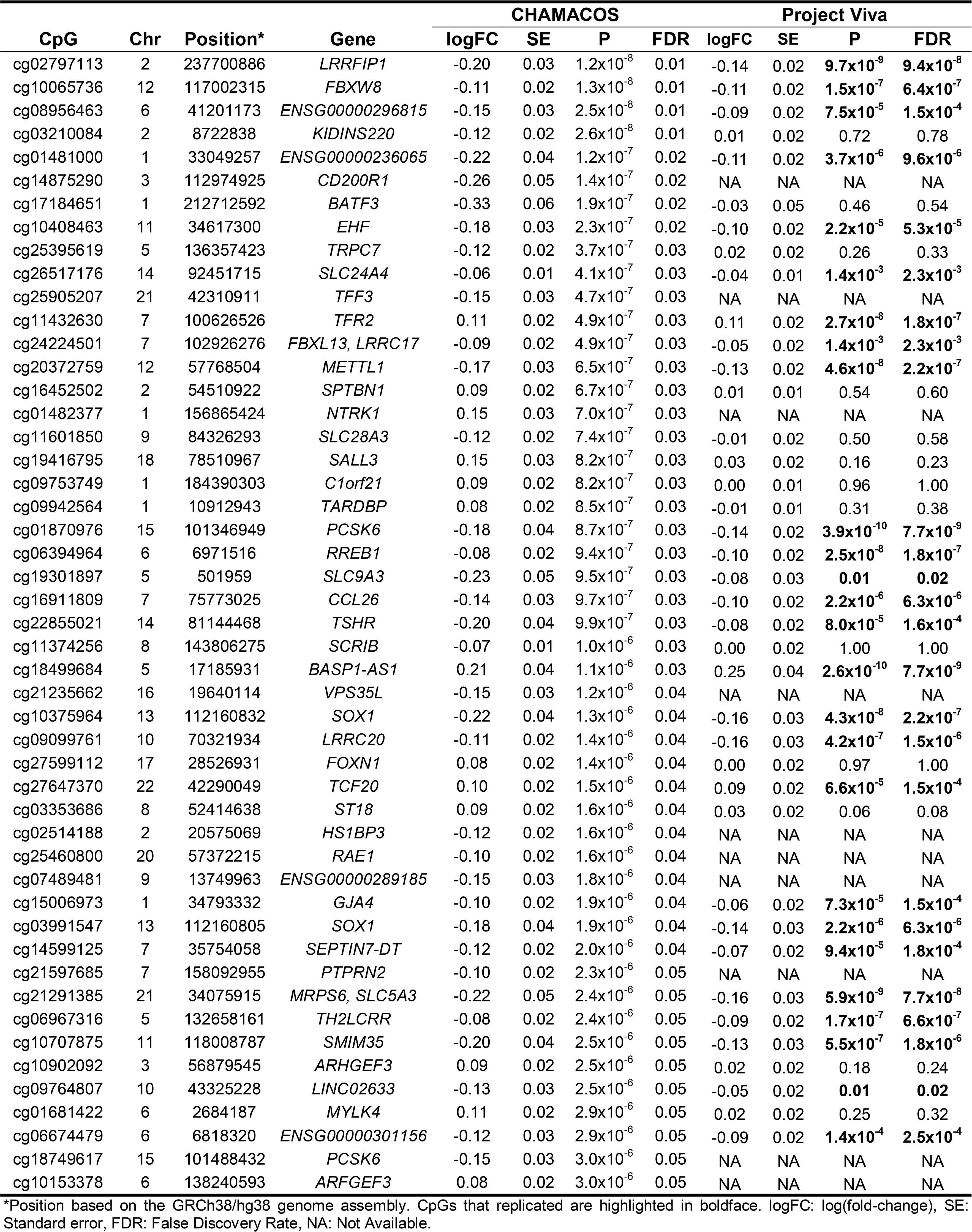
Summary table of CpGs significantly associated with FeNO.

| CpG | Chr | Position* | Gene | CHAMACOS |  |  |  | Project Viva |  |  |  |
| --- | --- | --- | --- | --- | --- | --- | --- | --- | --- | --- | --- |
|  |  |  |  | logFC | SE | P | FDR | logFC | SE | P | FDR |
| cg02797113 | 2 | 237700886 | LRRFIP1 | -0.20 | 0.03 | 1.2x10<sup>-8</sup> | 0.01 | -0.14 | 0.02 | **9.7x10<sup>-9</sup>** | **9.4x10<sup>-8</sup>** |
| cg10065736 | 12 | 117002315 | FBXW8 | -0.11 | 0.02 | 1.3x10<sup>-8</sup> | 0.01 | -0.11 | 0.02 | **1.5x10<sup>-7</sup>** | **6.4x10<sup>-7</sup>** |
| cg08956463 | 6 | 41201173 | ENSG00000296815 | -0.15 | 0.03 | 2.5x10<sup>-8</sup> | 0.01 | -0.09 | 0.02 | **7.5x10<sup>-5</sup>** | **1.5x10<sup>-4</sup>** |
| cg03210084 | 2 | 8722838 | KIDINS220 | -0.12 | 0.02 | 2.6x10<sup>-8</sup> | 0.01 | 0.01 | 0.02 | 0.72 | 0.78 |
| cg01481000 | 1 | 33049257 | ENSG00000236065 | -0.22 | 0.04 | 1.2x10<sup>-7</sup> | 0.02 | -0.11 | 0.02 | **3.7x10<sup>-6</sup>** | **9.6x10<sup>-6</sup>** |
| cg14875290 | 3 | 112974925 | CD200R1 | -0.26 | 0.05 | 1.4x10<sup>-7</sup> | 0.02 | NA | NA | NA | NA |
| cg17184651 | 1 | 212712592 | BATF3 | -0.33 | 0.06 | 1.9x10<sup>-7</sup> | 0.02 | -0.03 | 0.05 | 0.46 | 0.54 |
| cg10408463 | 11 | 34617300 | EHF | -0.18 | 0.03 | 2.3x10<sup>-7</sup> | 0.02 | -0.10 | 0.02 | **2.2x10<sup>-5</sup>** | **5.3x10<sup>-5</sup>** |
| cg25395619 | 5 | 136357423 | TRPC7 | -0.12 | 0.02 | 3.7x10<sup>-7</sup> | 0.03 | 0.02 | 0.02 | 0.26 | 0.33 |
| cg26517176 | 14 | 92451715 | SLC24A4 | -0.06 | 0.01 | 4.1x10<sup>-7</sup> | 0.03 | -0.04 | 0.01 | **1.4x10<sup>-3</sup>** | **2.3x10<sup>-3</sup>** |
| cg25905207 | 21 | 42310911 | TFF3 | -0.15 | 0.03 | 4.7x10<sup>-7</sup> | 0.03 | NA | NA | NA | NA |
| cg11432630 | 7 | 100626526 | TFR2 | 0.11 | 0.02 | 4.9x10<sup>-7</sup> | 0.03 | 0.11 | 0.02 | **2.7x10<sup>-8</sup>** | **1.8x10<sup>-7</sup>** |
| cg24224501 | 7 | 102926276 | FBXL13, LRRC17 | -0.09 | 0.02 | 4.9x10<sup>-7</sup> | 0.03 | -0.05 | 0.02 | **1.4x10<sup>-3</sup>** | **2.3x10<sup>-3</sup>** |
| cg20372759 | 12 | 57768504 | METTL1 | -0.17 | 0.03 | 6.5x10<sup>-7</sup> | 0.03 | -0.13 | 0.02 | **4.6x10<sup>-8</sup>** | **2.2x10<sup>-7</sup>** |
| cg16452502 | 2 | 54510922 | SPTBN1 | 0.09 | 0.02 | 6.7x10<sup>-7</sup> | 0.03 | 0.01 | 0.01 | 0.54 | 0.60 |
| cg01482377 | 1 | 156865424 | NTRK1 | 0.15 | 0.03 | 7.0x10<sup>-7</sup> | 0.03 | NA | NA | NA | NA |
| cg11601850 | 9 | 84326293 | SLC28A3 | -0.12 | 0.02 | 7.4x10<sup>-7</sup> | 0.03 | -0.01 | 0.02 | 0.50 | 0.58 |
| cg19416795 | 18 | 78510967 | SALL3 | 0.15 | 0.03 | 8.2x10<sup>-7</sup> | 0.03 | 0.03 | 0.02 | 0.16 | 0.23 |
| cg09753749 | 1 | 184390303 | C1orf21 | 0.09 | 0.02 | 8.2x10<sup>-7</sup> | 0.03 | 0.00 | 0.01 | 0.96 | 1.00 |
| cg09942564 | 1 | 10912943 | TARDBP | 0.08 | 0.02 | 8.5x10<sup>-7</sup> | 0.03 | -0.01 | 0.01 | 0.31 | 0.38 |
| cg01870976 | 15 | 101346949 | PCSK6 | -0.18 | 0.04 | 8.7x10<sup>-7</sup> | 0.03 | -0.14 | 0.02 | **3.9x10<sup>-10</sup>** | **7.7x10<sup>-9</sup>** |
| cg06394964 | 6 | 6971516 | RREB1 | -0.08 | 0.02 | 9.4x10<sup>-7</sup> | 0.03 | -0.10 | 0.02 | **2.5x10<sup>-8</sup>** | **1.8x10<sup>-7</sup>** |
| cg19301897 | 5 | 501959 | SLC9A3 | -0.23 | 0.05 | 9.5x10<sup>-7</sup> | 0.03 | -0.08 | 0.03 | **0.01** | **0.02** |
| cg16911809 | 7 | 75773025 | CCL26 | -0.14 | 0.03 | 9.7x10<sup>-7</sup> | 0.03 | -0.10 | 0.02 | **2.2x10<sup>-6</sup>** | **6.3x10<sup>-6</sup>** |
| cg22855021 | 14 | 81144468 | TSHR | -0.20 | 0.04 | 9.9x10<sup>-7</sup> | 0.03 | -0.08 | 0.02 | **8.0x10<sup>-5</sup>** | **1.6x10<sup>-4</sup>** |
| cg11374256 | 8 | 143806275 | SCRIB | -0.07 | 0.01 | 1.0x10<sup>-6</sup> | 0.03 | 0.00 | 0.02 | 1.00 | 1.00 |
| cg18499684 | 5 | 17185931 | BASP1-AS1 | 0.21 | 0.04 | 1.1x10<sup>-6</sup> | 0.03 | 0.25 | 0.04 | **2.6x10<sup>-10</sup>** | **7.7x10<sup>-9</sup>** |
| cg21235662 | 16 | 19640114 | VPS35L | -0.15 | 0.03 | 1.2x10<sup>-6</sup> | 0.04 | NA | NA | NA | NA |
| cg10375964 | 13 | 112160832 | SOX1 | -0.22 | 0.04 | 1.3x10<sup>-6</sup> | 0.04 | -0.16 | 0.03 | **4.3x10<sup>-8</sup>** | **2.2x10<sup>-7</sup>** |
| cg09099761 | 10 | 70321934 | LRRC20 | -0.11 | 0.02 | 1.4x10<sup>-6</sup> | 0.04 | -0.16 | 0.03 | **4.2x10<sup>-7</sup>** | **1.5x10<sup>-6</sup>** |
| cg27599112 | 17 | 28526931 | FOXN1 | 0.08 | 0.02 | 1.4x10<sup>-6</sup> | 0.04 | 0.00 | 0.02 | 0.97 | 1.00 |
| cg27647370 | 22 | 42290049 | TCF20 | 0.10 | 0.02 | 1.5x10<sup>-6</sup> | 0.04 | 0.09 | 0.02 | **6.6x10<sup>-5</sup>** | **1.5x10<sup>-4</sup>** |
| cg03353686 | 8 | 52414638 | ST18 | 0.09 | 0.02 | 1.6x10<sup>-6</sup> | 0.04 | 0.03 | 0.02 | 0.06 | 0.08 |
| cg02514188 | 2 | 20575069 | HS1BP3 | -0.12 | 0.02 | 1.6x10<sup>-6</sup> | 0.04 | NA | NA | NA | NA |
| cg25460800 | 20 | 57372215 | RAE1 | -0.10 | 0.02 | 1.6x10<sup>-6</sup> | 0.04 | NA | NA | NA | NA |
| cg07489481 | 9 | 13749963 | ENSG00000289185 | -0.15 | 0.03 | 1.8x10<sup>-6</sup> | 0.04 | NA | NA | NA | NA |
| cg15006973 | 1 | 34793332 | GJA4 | -0.10 | 0.02 | 1.9x10<sup>-6</sup> | 0.04 | -0.06 | 0.02 | **7.3x10<sup>-5</sup>** | **1.5x10<sup>-4</sup>** |
| cg03991547 | 13 | 112160805 | SOX1 | -0.18 | 0.04 | 1.9x10<sup>-6</sup> | 0.04 | -0.14 | 0.03 | **2.2x10<sup>-6</sup>** | **6.3x10<sup>-6</sup>** |
| cg14599125 | 7 | 35754058 | SEPTIN7-DT | -0.12 | 0.02 | 2.0x10<sup>-6</sup> | 0.04 | -0.07 | 0.02 | **9.4x10<sup>-5</sup>** | **1.8x10<sup>-4</sup>** |
| cg21597685 | 7 | 158092955 | PTPRN2 | -0.10 | 0.02 | 2.3x10<sup>-6</sup> | 0.05 | NA | NA | NA | NA |
| cg21291385 | 21 | 34075915 | MRPS6, SLC5A3 | -0.22 | 0.05 | 2.4x10<sup>-6</sup> | 0.05 | -0.16 | 0.03 | **5.9x10<sup>-9</sup>** | **7.7x10<sup>-8</sup>** |
| cg06967316 | 5 | 132658161 | TH2LCRR | -0.08 | 0.02 | 2.4x10<sup>-6</sup> | 0.05 | -0.09 | 0.02 | **1.7x10<sup>-7</sup>** | **6.6x10<sup>-7</sup>** |
| cg10707875 | 11 | 118008787 | SMIM35 | -0.20 | 0.04 | 2.5x10<sup>-6</sup> | 0.05 | -0.13 | 0.03 | **5.5x10<sup>-7</sup>** | **1.8x10<sup>-6</sup>** |
| cg10902092 | 3 | 56879545 | ARHGEF3 | 0.09 | 0.02 | 2.5x10<sup>-6</sup> | 0.05 | 0.02 | 0.02 | 0.18 | 0.24 |
| cg09764807 | 10 | 43325228 | LINC02633 | -0.13 | 0.03 | 2.5x10<sup>-6</sup> | 0.05 | -0.05 | 0.02 | **0.01** | **0.02** |
| cg01681422 | 6 | 2684187 | MYLK4 | 0.11 | 0.02 | 2.9x10<sup>-6</sup> | 0.05 | 0.02 | 0.02 | 0.25 | 0.32 |
| cg06674479 | 6 | 6818320 | ENSG00000301156 | -0.12 | 0.03 | 2.9x10<sup>-6</sup> | 0.05 | -0.09 | 0.02 | **1.4x10<sup>-4</sup>** | **2.5x10<sup>-4</sup>** |
| cg18749617 | 15 | 101488432 | PCSK6 | -0.15 | 0.03 | 3.0x10<sup>-6</sup> | 0.05 | NA | NA | NA | NA |
| cg10153378 | 6 | 138240593 | ARFGEF3 | 0.08 | 0.02 | 3.0x10<sup>-6</sup> | 0.05 | NA | NA | NA | NA |

\*Position based on the GRCh38/hg38 genome assembly. CpGs that replicated are highlighted in boldface. logFC: log(fold-change), SE: Standard error, FDR: False Discovery Rate, NA: Not Available.

### Differentially methylated regions

A total of 557 DMRs were associated with respiratory outcomes, including asthma, FeNO, allergic sensitization, and lung function parameters (FDR<0.05, **Figure 2**, **Table S2**). Most of the associations were reported for FeNO (327 DMRs), FEF_25-75_ (111 DMRs), and FEV_1_/FVC (86 DMRs). The most significant DMRs for each outcome included the genes *ATF6B* associated with FEF_25-75_, *IRX1* with FeNO, *SPG7* with allergic sensitization, *KIF26A* with FEV_1_/FVC, *MBP* with PEF, *GGTLC1* with asthma (ever), *MGME1* with late-onset asthma, *EEFSEC* with FEV_1_, *NRP2* with current asthma, *ARGLU1* with early-onset asthma, and *RAB19* with FVC. We also detected genes related to eosinophilic inflammation (*EPX*), cytokine and chemokine pathways (*IL13*, *IL1R1*, *IL31RA*, *ILDR1*, *TOLLIP*, *TNF*, *TNFRSF19*, *TNFAIP6*, *TNFSF4*, *CCR6*), T2 inflammation and T-cell activity (*GATA3*, *RUNX3*, *BACH2*), airway inflammation (*DUSP6*, *THBS1, TREML2*), and immune cell regulation (*JAK3*, *ETS1*).

**Figure 2.**
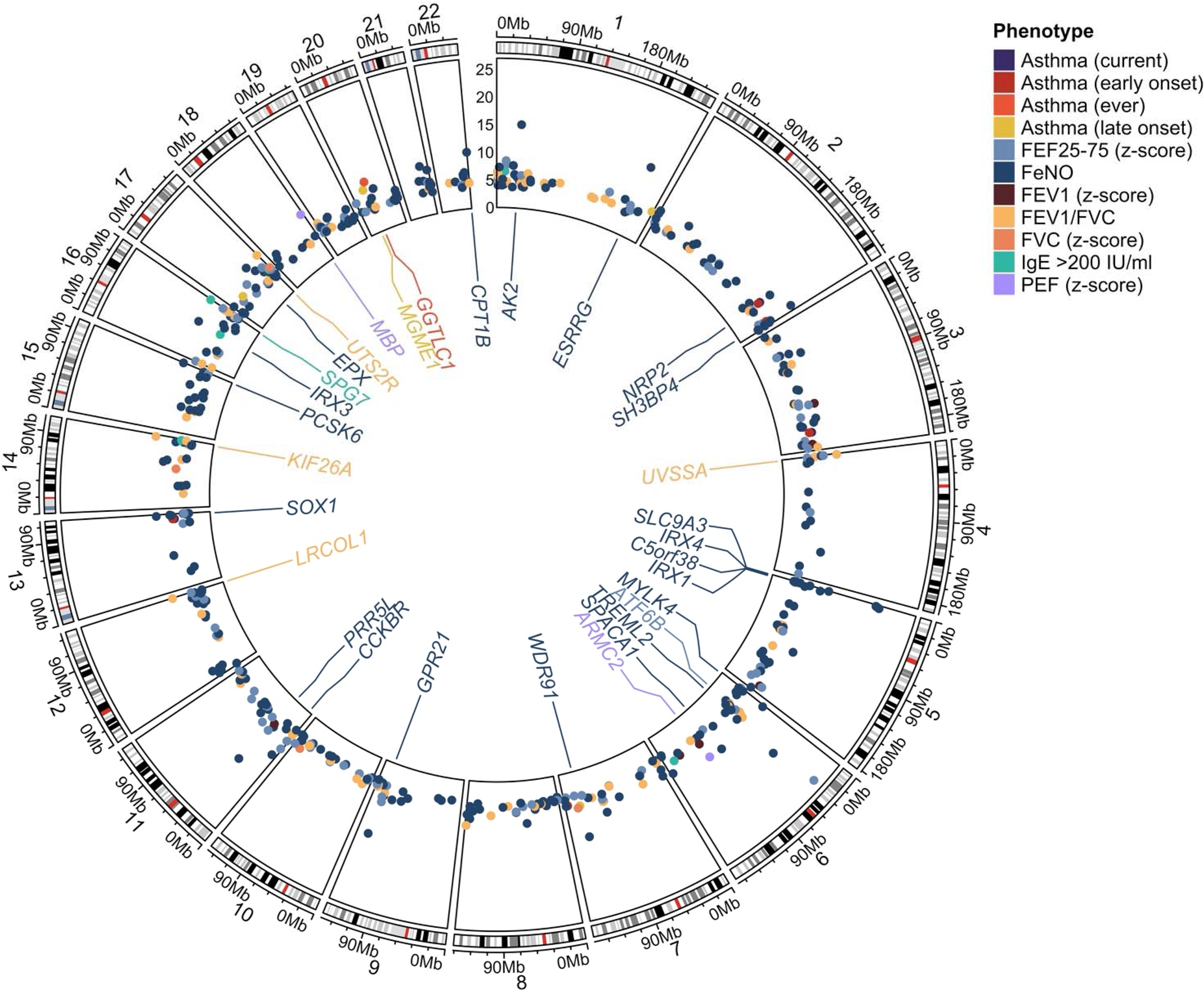
Circos plot displaying differentially methylated regions (DMRs) associated with respiratory phenotypes. Each point corresponds to one DMR, with the radial (y) axis showing the association -log_10_(*p*). Points are colored by phenotype. Gene annotations are provided for the top three DMRs per phenotype.

### Replication analyses

We followed up identified CpGs and DMRs for replication in 540 adolescents from Project Viva (**Table 1**). Briefly, the mean (SD) age was 13.0 (0.7) years and 49.1% were female. Most were non-Hispanic White (65.9%), 64 (13.9%) had current asthma, 84 (23.3%) had allergic sensitization, and the mean (SD) FeNO was 27.6 ppb (29.4).

Among the 49 CpGs associated with FeNO in the discovery phase, 39 were available in Project Viva, and 26 (66%) were replicated with the same effect direction (*p*<0.05, **Table 3**, **Figure 3A**). The CpG with the most significant replication signal was cg01870976, annotated to *BASP1-AS1* (logFC=0.25, *p*=2.6×10^−10^). We also reported replication for CpGs annotated to genes associated with inflammation (*RREB1*, *GJA4*), TNF signaling (*LRRFIP1*), eosinophil recruitment (*CCL26*), epithelial integrity (*EHF*, *SLC9A3*), T2 inflammation (*TH2LCRR*), thyroid hormone signaling (*TSHR*), bronchodilator response (*SLC24A4*), and atopy (*SLC9A3*).

**Figure 3.**
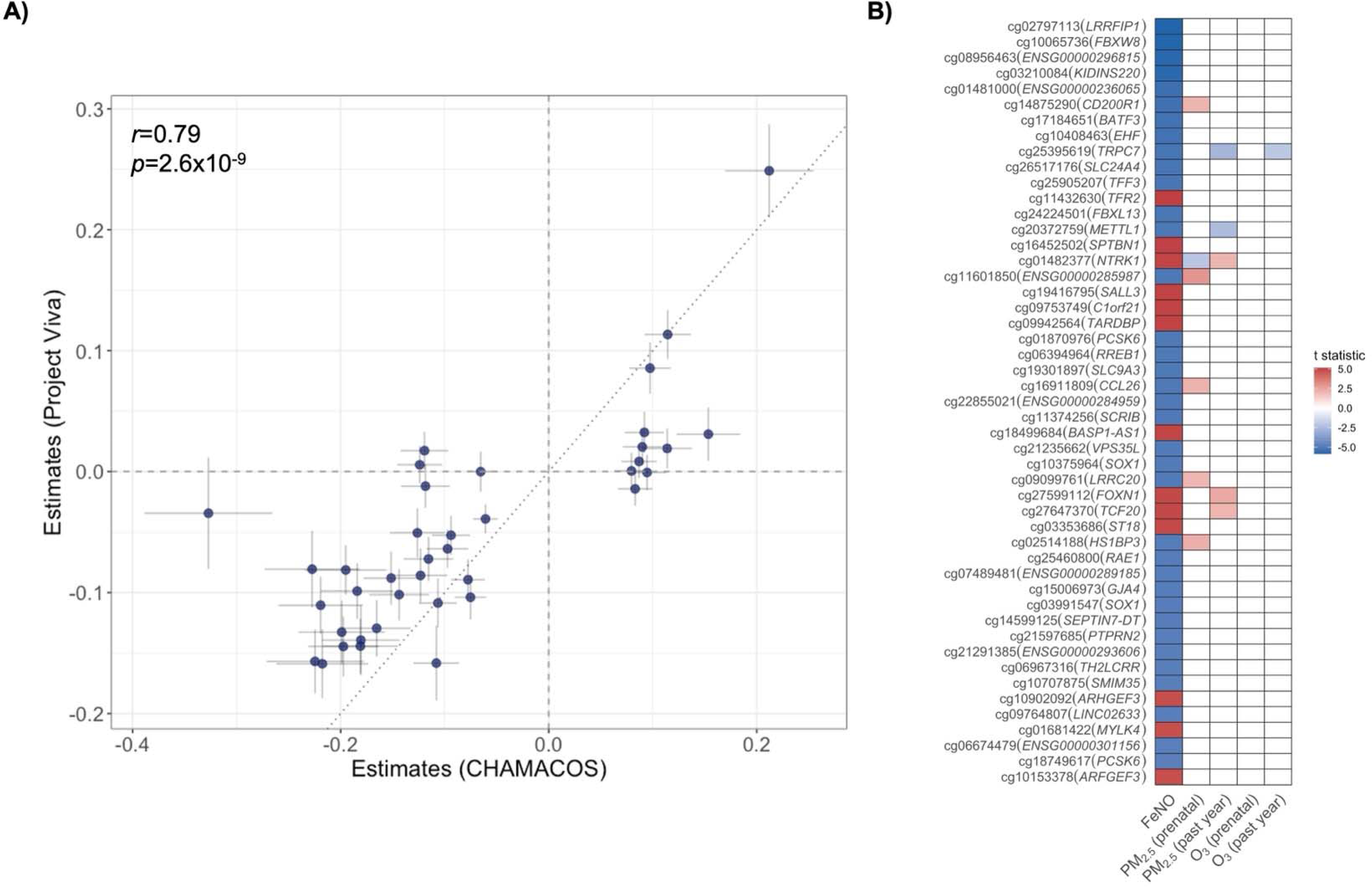
**A)** Scatter plot showing the estimates in CHAMACOS (x-axis) and Project Viva (y-axis) of FeNO-associated CpGs followed up for replication. Error bars are also represented. A high correlation was observed among estimates based on Pearson’s correlation coefficient. **B)** Heatmap of the association of FeNO-related CpGs with air pollution exposures to PM_2.5_ and O_3_ during the prenatal period and in the past year. CpGs significantly associated with FeNO are represented on the x-axis. CpGs positively associated with air pollution are colored red, and those negatively associated are colored blue. Only associations at *p*<0.05 are colored.

Among the 369 DMRs available for replication in Project Viva, 103 were replicated with the same effect direction for the same outcomes as in the discovery (*p*<0.05, **Table S3**). These included 97 DMRs associated with FeNO, five with FEV_1_/FVC (*STAR*, *LRCOL1*, *TMEM250, PIP4K2A*, *MEF2D*), and one (*ARGLU1*) with current asthma. Replicated DMRs associated with FeNO included asthma-related genetic loci such as *EPX*, *IL13*, *TOLLIP*, *TNF*, *TNFRSF19*, *CCR6, THBS1, MYLK4, SLC9A3,* and *TREML2*.

### Association of CpGs with air pollution

Among the 49 individual CpGs significantly associated with FeNO, 10 of them were nominally associated with PM_2.5_ exposures during early life (prenatal) and/or in the past year in CHAMACOS (*p*<0.05, **Table S4**). CpGs associated with prenatal exposures showed effect directions opposite to those observed for the association with FeNO, whereas those associated with exposures in the past year were in the same direction as FeNO (**Figure 3B**). Only one CpG was associated with O_3_ exposure in the past year. However, no association remained significant after multiple comparisons adjustment (FDR>0.05).

### In silico associations with gene expression and genetic variation

A total of 15 CpGs associated with FeNO in our study were previously reported as cis-eQTM of 52 genes in nasal epithelia (**Figure S2**, **Table S5**), including genes involved in T-cell activity (*LCK*), airway epithelial integrity (*FOXP4*), airway inflammation (*TREM2*, *HDAC1*, *LRCH4, GJA4*), innate immunity (*TRIM62*), and atopy (*SLC9A3*). We also detected two CpGs acting as cis-eQTMs of *PCSK6*, including one of the most significant replication signals. Additionally, nasal DNAm at only six CpGs associated with FeNO were previously reported to be regulated by meQTLs (**Table S6**).

### Enrichment analyses

Among gene annotations of CpGs where nasal DNAm was suggestively associated with respiratory outcomes (FDR<0.2), we found enrichment of genes linked to multiple traits, including obesity-related traits and longevity, as well as genes regulated by NFKB1 and NR3C1 (glucocorticoid receptor) in bronchial epithelial cells (FDR<0.05, **Figure 4**, **Table S7**). The gene-set enrichment analysis based on DMRs showed an overrepresentation of genes associated with asthma-related traits (*i.e.,* asthma, allergic diseases, IgE levels, and lung function), comorbidities and risk factors (i.e., smoking and obesity-related traits), candidate drugs for asthma treatment (*i.e.,* trichostatin A and metformin), and biological pathways such as PI3K-AKT, MAPK, Estrogen, TNF-α (via NF-kB), and IL-2/STAT5 signaling pathways (FDR<0.05, **Table S8**).

**Figure 4.**
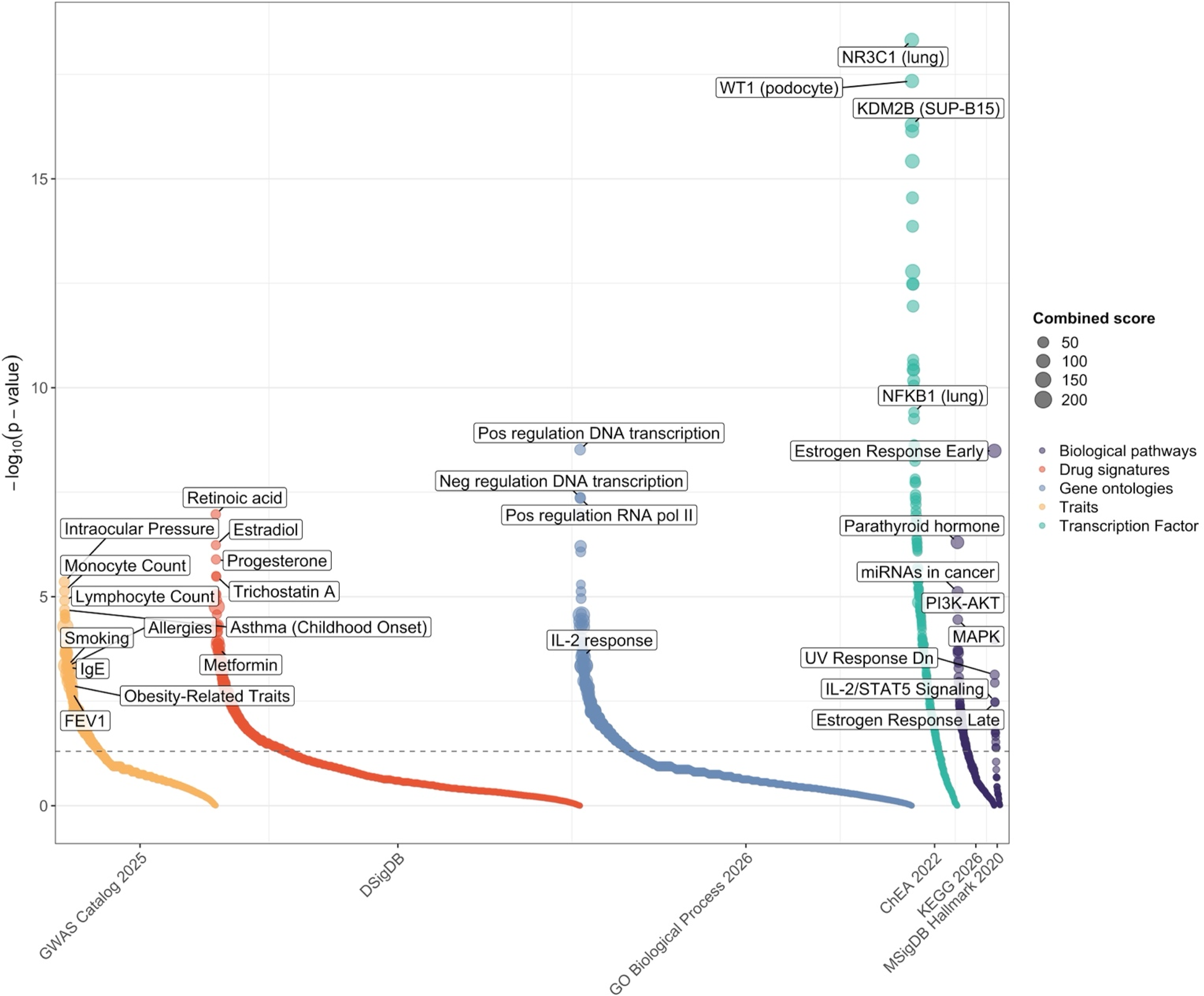
Manhattan-like plot of enrichment terms based on differentially methylated regions (DMRs). Each point represents a term, grouped along the x-axis by database. The y-axis shows statistical significance of enrichment analysis as -log_10_(p-value). Point color indicates the category of the terms (purple: biological pathways, red: drug signatures, purple: gene ontologies, yellow: human traits, turquoise: transcription factor), and point size is proportional to the combined score (combination of p-values and odds ratio calculated as score=-log(p)* odds ratio). Annotation is provided for the top three most significant terms for each database and selected terms of interest for asthma and allergies. The dashed horizontal line represents *p*<0.05.

### Epigenetic clocks

The Skin&Blood clock estimated from nasal DNAm data was moderately correlated with chronological age in CHAMACOS (r=0.31, *p*=1.6×10^−9^), while a marginal correlation was observed for HorvathAge (r=0.10, *p*=0.053) (**Figure S3**). We observed accelerated epigenetic aging associated with higher FeNO (HorvathAge: β=1.6 years, *p*=1.3×10^−6^; SkinBloodAge: β=0.3 years, *p*=0.008) and allergic sensitization (HorvathAge: β=2.4 years, *p*=0.002) (**Table S9**). In the sensitivity analysis adjusted for cell-type heterogeneity, only the association between FeNO and higher SkinBloodAge remained significant (β=0.26 years, *p*=0.046, **Table S9**).

## DISCUSSION

To the best of our knowledge, this is the first nasal EWAS comprehensively analyzing multiple respiratory outcomes in Mexican American young adults. We identified 49 CpGs and 557 DMRs associated with respiratory outcomes, of which 39 CpGs and 102 DMRs replicated in adolescents from a younger and multi-ethnic population (66% White). Most of the signals were associated with FeNO, with a few associated with current asthma and lung function. Fifteen FeNO-related CpGs were *in silico* associated with nasal epithelial expression of genes involved in airway epithelial integrity, inflammation, and immunity. We observed an enrichment of genes associated with respiratory and allergic outcomes, longevity, asthma risk factors (obesity and smoking), and several biological pathways (*e.g.,* PI3K-AKT, TNF-α, and IL-2), as well as genes regulated by the GR and NF-kB transcription factors and candidate asthma drugs. Finally, we found an accelerated epigenetic aging associated with higher FeNO and atopy.

Similar to a previous nasal EWAS of asthma and airway inflammation biomarkers in children (6), FeNO was the biomarker most strongly associated with the nasal epigenome in our study. Although we did not find a robust DNAm signature for asthma and the other evaluated respiratory outcomes, we found an enrichment of CpGs located near annotated genes previously linked to asthma, allergies, lung function, and IgE, supporting the link between our findings and respiratory health. We hypothesized that lack of signals could relate to reduced statistical power due to the relatively low number of asthma cases in a pre-birth cohort, or that signals in young adults may be weaker than in children.

Supporting the plausibility of our findings, we detected numerous epigenetic markers involved in pathways significant for respiratory health, such as inflammation (*RREB1*, *GJA4, IL13, CCR6, THBS1*), epithelial integrity (*EHF*, *SLC9A3*), eosinophil activity (*EPX*, *CCL26*), T2 inflammation (*TH2LCRR, TREML2*), and smooth muscle tone regulation (*MYLK4*). For example, *EPX* (eosinophil peroxidase) is one of the most robust EWAS signals associated with asthma in different tissues, which participates in host defense and interleukin regulation (30). Multiple solute carriers and intracellular transport genes have been widely associated with asthma and inflammatory biomarkers, with *SLC9A3* DNAm associated with atopy and expression of genes linked to asthma risk (10,31). The myosin light chain kinase (*MYLK*) family is related to asthma and exacerbation risk given their role in muscle contraction and inflammation (32), and although little is known about the role of *MYLK4*, DNAm at this gene potentially underlies the mechanism of action of albuterol (33).

Our top hit in CHAMACOS was the CpG cg02797113 associated with FeNO and annotated to *LRRFIP1* (LRR binding FLII interacting protein 1). *LRRFIP1* contributes to host defense against microbial infections via interferon-β (IFN-β) production and regulates TNF-α expression (34,35). Notably, we detected an enrichment of epigenetic loci implicated in TNF-α pathway associated with FeNO and lung function (*TNF*, *TNFRSF19*, *TNFAIP6*, *TNFSF4*). TNF-α is a cytokine and a candidate target for asthma treatment that plays a key role in host defense and inflammation homeostasis (9). Furthermore, the CpG cg01870976, annotated to *PCSK6* (proprotein Convertase Subtilisin/Kexin Type 6), showed one of the most significant replication signals and was *in silico* associated with *PCSK6* nasal expression (28). *PCSK6* encodes a calcium-dependent serine protease highly expressed in airway epithelia that participates in the TGF-β, NF-κB, IL-1, and IL-6 inflammatory pathways and metalloprotease activity (36–38). Nasal DNAm levels at this CpG have been associated with atopy, asthma, and rhinitis (6,10,39).

Furthermore, we detected epigenetic loci participating in the phosphatidylinositol 3-kinase (PI3K), mitogen-activated protein kinase (MAPK), and estrogen signaling pathways. Both PI3K and MAPK pathways have been extensively associated with asthma pathology, airway inflammation and infections, and poor steroid response (40–42). Enrichment in estrogen response may suggest sex-specific effects of detected DNAm markers on airway inflammation (43). Beyond established asthma-related pathways, we identified eight CpGs newly associated with FeNO that we could not replicate because they are unique to the InfiniumMethylationEPICv2 microarray. These epigenetic loci remain of interest for respiratory health given prior links to airway inflammation (*CD200R1*) (44), interleukin production (*NTRK1*) (45), T2 asthma (*TFF3*) (46), and DNAm associations with childhood asthma and stress (47) by EWAS or in vitro studies.

Additionally, among suggestive signals from EWAS, we found an enrichment of genes involved in obesity and likely regulated by the antidiabetic drug metformin. Obesity is a major risk factor for asthma, exacerbations, and poor response to therapies both in children and adults (48). Genetic and nasal transcriptomic studies have identified specific biological pathways underlying both conditions, supporting a shared molecular basis (49,50). After adjustment for BMI, our findings support DNAm as a molecular modification underlying both conditions and suggest the potential repurposing of metformin for asthma treatment (51,52). We also observed enrichment of genes putatively regulated by trichostatin A and tanespimycin, two compounds primarily investigated as anticancer therapies that have shown airway anti-inflammatory effects in animal and in vitro models of asthma (53,54). Notably, enrichment for trichostatin A and inflammatory transcriptional regulators (NF-kB and GR) has been reported by independent epigenetic and microbiome studies of asthma traits (9,33,55,56).

Moreover, we observed enrichment of genetic loci related to smoking and longevity. Smoking is a preventable risk factor for all-cause mortality and disability. Substantial evidence indicates that smoking induces DNAm signatures (e.g., *AHRR*) that can persist into childhood and affect genes relevant to asthma (7). Because our analyses were adjusted for recent smoking, these findings suggest that smoking-induced DNAm changes may influence airway inflammation in young adults. In addition to smoking, air pollution is a major environmental exposure contributing to the onset and severity of respiratory diseases (7). A recent study suggested that blood DNAm may mediate the effect of air pollution on asthma control (11). We found that some FeNO-associated signals were also associated with PM_2.5_ exposure, implicating inflammatory genes (*NTRK1, TRPC7*) (45,57) and a locus previously linked to asthma and atopy in EWAS using airway samples (*METTL1*) (6,10,38). Collectively, these results support nasal DNAm as a potential link between smoking, air pollution, and respiratory health, warranting further investigation in future studies.

Finally, aging substantially contributes to the pathogenesis of respiratory diseases, and accelerated epigenetic aging has been associated with poorer respiratory health (58,59) and with risk factors such as smoking (60,61). We also found that participants with higher FeNO and elevated IgE levels had greater epigenetic age acceleration, consistent with prior findings in children from Project Viva (6). These findings underscore the potential utility of epigenetic clocks in the management of respiratory diseases. It is also important to note that the CHAMACOS and Project Viva datasets differed substantially in participant age and genetic ancestry, which may limit the transferability of the findings. Nevertheless, the consistency observed for several epigenetic markers supports the robustness of these findings.

Our study has several strengths. First, we analyzed two pre-birth cohorts deeply characterized for multiple respiratory and allergic outcomes, as well as for air pollution exposure using highly accurate and validated models. Second, we analyzed nasal samples mostly composed of epithelial cells, which are surrogates of lower airways and the first line of defense of the airways, in Mexican Americans, a population reporting high asthma burden despite low prevalence (4). Third, we analyzed >800,000 CpGs using the most comprehensive Illumina Infinium MethylationEPICv2 microarray released to date. Fourth, we applied state-of-the-art analytical methods to ensure the robustness of our findings, including a stringent QC of microarray data, confounders and multiple comparisons adjustment, and replication in an independent population.

However, several limitations should be acknowledged. First, because we analyzed cohorts recruited pre-birth, the number of asthma/allergic cases was relatively small, which may reduce statistical power and contribute to null findings for some outcomes. Second, the cross-sectional design did not infer causality, and longitudinal approaches would be needed to address temporality. Third, we could not directly measure nasal cell type composition, although we applied a reference-free method to mitigate this confounder. Fourth, due to the absence of RNA-seq and genotyping data for study participants, we conducted *in silico* QTL analyses. Fifth, although a substantial proportion of DNAm signals were replicated, replication may have been limited by heterogeneity in age groups and ancestry across populations and by the unavailability of certain spirometry parameters in the replication dataset.

## CONCLUSIONS

We identified multiple CpGs and DMRs, of which nasal DNAm levels were predominantly associated with FeNO in young adult Mexican Americans. Many of the FeNO-related signals were replicated in multi-ethnic adolescents. These markers likely participate in airway inflammation, epithelial barrier integrity, eosinophil recruitment, and immune responses. Higher FeNO was also associated with greater nasal epigenetic age. Genes associated with respiratory health were enriched for allergic and respiratory traits, asthma comorbidities, longevity, cytokine signaling pathways, and gene signatures of candidate drugs for asthma treatment. In contrast, in this study, we found limited evidence for associations between the nasal methylome and asthma, IgE, and lung function.

## ACKNOWLEDGMENTS

The authors thank all the participants (mothers and offspring) from the CHAMACOS and Project Viva birth cohorts, as well as all researchers, clinicians, and people involved in the establishment of these cohorts.

## AUTHOR CONTRIBUTIONS

JP-G, RBG, and AC were involved in the conceptualization and design of the study; JP-G and RBG in data curation and formal analyses; BE, JB, KK, WG, SLR-S, M-FH, EO, DRG, SH, NH, and RBG in study conceptualization, sample collection and processing, cohort management, and data analysis and interpretation; JP-G in visualization and writing of the original draft; RBG and AC in funding acquisition and project supervision. All authors were involved in the critical revision of the manuscript and have read and agreed to the published version of the manuscript. The authors agree to be accountable for all aspects of the work, ensuring its scientific accuracy and integrity.

## DATA AVAILABILITY

The summary statistics of all association analyses supporting the main conclusions of this study are reported in the main text and supplementary material. We additionally uploaded the full summary statistics of EWAS from the discovery phase to the Zenodo repository (DOI: 10.5281/zenodo.23699726), which will be publicly available after manuscript acceptance. Data reported in this study are available from the corresponding author upon reasonable request and with IRB approval. To access CHAMACOS data or samples, investigators must submit an Initial Data Use Application (IDUA) to the Study Coordinator, Katherine R. Kogut. The CHAMACOS Executive Committee reviews applications. Detailed protocols and application instructions are available at https://cerch.berkeley.edu/investigators. In the case of Project Viva, data included in this manuscript are not publicly available because Project Viva’s historic consents did not allow for public data sharing. In accordance with Project Viva policies, datasets are available upon reasonable request via the Project Viva ROADMaP portal. Please see https://www.projectviva.org/for-investigators for information about how to request data. All data collection instruments are also available via the ROADMaP.

## ABBREVIATIONS

BMI: Body mass index
CHAMACOS: Center for the Health Assessment of Mothers and Children of Salinas
CpG sites: 5’-cytosine-phosphate-guanine-3’ dinucleotides
DMRs: Differentially methylated regions
DNAm: DNA methylation
eQTM: Expression quantitative trait methylation
EPA: Environmental Protection Agency
EWAS: Epigenome-wide association studies
FDR: False discovery rate
FEF_25-75_: Forced expiratory flow between 25% and 75%
FeNO: Fractional exhaled nitric oxide
FEV_1_: Forced expiratory volume in one second
FVC: Forced vital capacity
GR: Glucocorticosteroid receptor (NR3C1)
GSEA: Gene-set enrichment analysis
IFN-β: Interferon-β
IgE: Immunoglobulin E
IL: Interleukin
MAPK: Mitogen-activated protein kinase
meQTL: Methylation quantitative trait loci
NF-kB: Nuclear factor kappa beta
PEF: Peak expiratory flow
PI3K: Phosphatidylinositol 3-kinase
TNF-α: Tumor necrosis factor-α

---

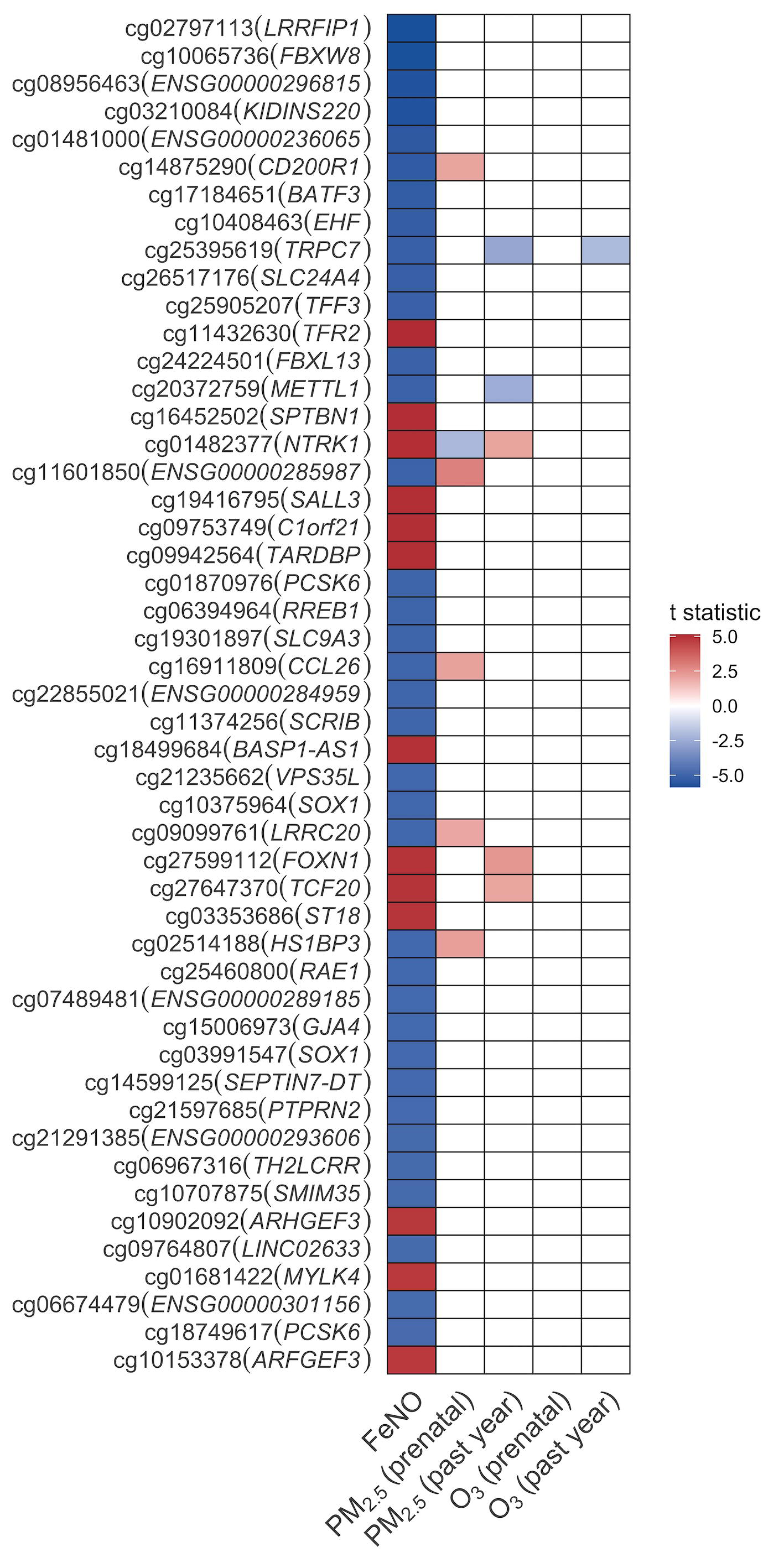

