## Supplementary Material for "Nasal Epigenome-Wide Association Study of Respiratory and Allergic Biomarkers in Mexican American Young Adults"

**SUPPLEMENTARY METHODS**

**Study Populations**

In the discovery phase, we analyzed 354 young adults from the Center for the Health Assessment of Mothers and Children of Salinas (CHAMACOS) cohort study (1,2). CHAMACOS is a longitudinal birth cohort study investigating the effects of prenatal and early postnatal environmental exposures on the health of children and young adults living in California’s Salinas Valley. This study was conducted in two phases: CHAM1 and CHAM2. In CHAM1, 601 pregnant women were recruited in the agricultural Salinas Valley, California, between 1999 and 2000 according to the following inclusion criteria: 1) ≥18 years old, 2) <20^th^ gestational week, 3) English- or Spanish-speaking, 4) eligible for low-income health insurance, and 4) planning to deliver at the county medical center. Among the 601 recruited women, 532 were followed during pregnancy, delivery, and their children were followed up at 1- to 2-year intervals. In CHAM2, mothers and 305 children were recruited when the child was 9 years old using the same inclusion criteria, completed a baseline data collection, and were followed up with the same study visits as CHAM1.

CHAMACOS children from CHAM 1 and 2 were eligible for the young adult respiratory study if they had completed a 16- or 18-year study visit (n=613; CHAM1 n=331; CHAM2=282). Of these, 440 completed the young adult respiratory questionnaire at approximately 23 years of age, 432 had valid spirometry data, and 409 had FeNO measurements. Trained research assistants collected nasal swabs from the anterior nares of 359 participants to yield respiratory epithelial cells using sterile cotton swabs placed in DNA lysis buffer after collection.

The University of California, Berkeley Committee for the Protection of Human Subjects approved all study activities. Written informed consent was obtained from mothers. Child verbal assent was obtained from children aged 7 and 9 years; written assent was obtained at age 14 years, and written consent was obtained at 18 years. This study followed the Strengthening the Reporting of Observational Studies in Epidemiology reporting guideline for cohort studies.

In the replication phase, we analyzed 540 nasal samples from adolescents (age 13) from the pre-birth cohort Project Viva, after removing 7 related individuals (siblings). A full description of the cohort design, nasal epigenome-wide DNAm profiling, and respiratory outcomes assessment was described elsewhere (3–5). Mothers provided written informed consent at recruitment and postpartum follow‐up visits. Participants provided consent for nasal swab collection, and study protocols were approved by the Institutional Review Board of Harvard Pilgrim Health Care.

**Respiratory Health and Biomarkers**

Lung function

Spirometry was conducted by a trained research assistant in accordance with ATS/ERS guidelines using the EasyOne® hand-held Diagnostic electronic spirometer (ndd, Chelmsford, MA) and EasyWare software (6). All participants were free of acute respiratory illness for 6 weeks before the spirometry. Parameters recorded from a minimum of 3 and a maximum of 8 maneuvers included the forced expiratory volume in one second (FEV_1_), forced vital capacity (FVC), the forced expiratory flow between 25% and 75% of FVC (FEF_25-75_), and the peak expiratory flow (PEF). Lung function parameters were normalized by age, sex, and height as Z-scores according to Global Lung Function Initiative (GLI) reference equations (7). We also estimated the ratio FEV_1_/FVC from raw data. A bronchodilator test was conducted to test for the presence of reversible airway obstruction.

Asthma

Asthma symptoms or medications in the past year was obtained via interview from the young adults and was defined as having asthma symptoms (experiencing wheezing, sleep disturbed by wheezing, speech limited by wheezing, wheezing during exercise, a cough without a cold, or a reported asthma attack in the past year (8); using medications (including asthma inhalers, as well as pills or other medicines used to treat asthma) in the past year; or ≥12% change in FEV_1_ after administration of albuterol during current spirometry testing, as a possible indicator of undiagnosed asthma. Asthma outcomes were created using maternal- and self-reported questionnaire data collected throughout participants’ lives from birth through 23 years of age. Individuals were classified as ever having asthma if either the mother or the young adult reported a physician diagnosis at any time. We defined asthma by age of onset as early (asthma diagnosed at 9 years old or earlier) or late (asthma diagnosed by 23 years old and after 9 years old). We defined ongoing asthma as having an asthma diagnosis at 9-years old or earlier and having asthma symptoms or medications in the past year.

Fractional exhaled nitric oxide (FeNO)

We measured FeNO levels using a portable electrochemical device (NIOX VERO®; Circassia Pharmaceuticals Inc., Morrisville, North Carolina) with a trained research assistant in accordance with established guidelines (9). Participants were instructed to inhale through a scrubbing filter and exhale into the FeNO analyzer for 10 seconds at a flow rate of 50 ml/s. The procedure was performed up to eight trials, and measurements from the two trials with values closest to each other were averaged. Results were expressed in parts per billion (ppb). In subsequent analyses, FeNO was log_2_ transformed to minimize skewed distributions.

Total immunoglobulin E (IgE) test

We used the Biopanda Total IgE Rapid Test (Biopanda Reagents, Belfast, UK) to detect human IgE antibodies in whole blood from fingerstick specimens following the manufacturer’s recommendations. A positive result (IgE levels ≥200 IU/ml) was used to define allergic sensitization.

**Nasal sampling**

Trained research assistants collected nasal swabs from the anterior nares to yield respiratory epithelial cells using sterile cotton swabs placed in DNA lysis buffer after collection. Nasal swabs were frozen until monthly shipping to the UC Berkeley Biorepository. Genomic DNA was isolated using the Maxwell 16 Buccal Swab LEV DNA Purification Kit according to the manufacturer’s instructions (Promega, Madison, WI, USA).

**Genome-wide DNAm measurement and quality control**

DNAm at 936,990 CpGs across the genome was measured in 372 nasal samples using the Infinium Illumina MethylationEPICv2 BeadChip (Illumina, San Diego, CA) following the manufacturer’s protocols. A standardized quality control (QC) of DNAm data was conducted using the *minfi*, *ENmix*, and *ewastools* R packages (10–12).

Bad-quality DNAm data points were defined by detection *p*>1×10^-6^ or detection beads <3. We filtered out low-quality probes (n=6,087) with ≥5% bad-quality data points across samples. Low-quality samples (n=0) were defined as having 1) ≥5% of bad-quality data points across probes, 2) total bisulfite intensity <3 standard deviations from bisulfite controls, and/or 3) outliers of bisulfite intensity or beta-value distribution. No outlier was detected for control probes included in the array for bisulfite conversion I and II, extension, hybridization, non-polymorphic, specificity I and II, and target removal. In addition, we inspected the log median intensities for methylated and unmethylated signals, and all samples clustered together, supporting no failed samples.

We corrected the background noise and dye bias using the out-of-band (oob) and the Regression on Logarithm of Internal Control (RELIC) method, respectively. We filtered out 176 probes with mapping inaccuracies and 46,338 with low-performance issues as described in the Illumina MethylationEPICv2 Manifest A1 (13). Inter-array intensity variation was corrected by quantile normalization, and the probe-type bias was adjusted using the Regression on Correlated Probes (RCP) method. Beta values were computed from bisulfite intensities, and outlier beta values (3*interquartile range rule) were set as missing data. We filtered out 7,472 CpGs with a high missingness rate (>5% missing data across samples). No samples showed a high missingness rate (>5% missing data across probes), and remaining missing data were imputed using the k-neighbor method.

We removed potential cross-sample contaminated samples, defined as 1) discordance between the reported sex and predicted sex based on sex chromosomes DNAm (n=1), and/or 2) mixed distribution of beta values for the 59 single nucleotide polymorphism genotyping control probes, outliers from the expected distributions for AA/AB/BB genotypes (n=1). Additionally, we filtered out these control probes and those annotated to sex chromosomes (Chr X and Y). Given that the Infinium MethylationEPICv2 array included a set of probes with 2-10 replicates, we filtered out 4,948 replicate probes according to a pre-defined list provided in the *MethylCall* R package (14). This list prioritizes probes 1) having the same sequence probe as in the EPICv1 array to improve reproducibility with previous studies, and 2) classified as superior probes. If the probe that should be retained based on the *MethylCall* R package was removed during the QC, we retained the other replicate or the one with the lowest root mean squared error when compared with whole-genome bisulfite sequencing (15).

Moreover, we filtered out potentially problematic probes, including those 1) without annotation in the GRCh38/hg38 build based on Illumina’s manifest, 2) with off-target binding sites (cross-reactive probes) (15,16), and 3) potentially polymorphic. We defined potentially polymorphic probes as those 1) flagged by the Illumina Manifest as SNP-CpG, SBE-CpG, or SNP-Probe with a minor allele frequency (MAF)≥5%, and showing a multimodal distribution, and/or 2) detected by the *MethyltoSNP* R function (17). Finally, we evaluated the correlation of DNAm for all probes among six pairs of technical-replicated samples and discarded 9 probes with a mean |Δ_Beta_|>0.2 among all replicates. The number of samples and CpGs that passed QC is summarized in **Supplementary Table S1**.

The beta-value matrix was corrected for batch effect (sample plate) while adjusting for major covariates using the ComBat-met algorithm and then log-transformed to M-values to minimize skewed distributions (18,19). All analyses were conducted on M-values.

**Air pollution exposure estimates**

We estimated PM_2.5_ (µg/m^3^) and O_3_ (ppb) exposure using two approaches. Ambient measurements from EPA monitoring stations in California were linked to the closest station to their geocoded residences within 50 km (20). If a station was missing monitoring data for a given month, the second closest station, within 50km, was used to replace the missing data. Any remaining missing measurements were replaced with the average value for the corresponding month and exposure from the monitoring station that was closest to the residence. Modeled air pollution exposures were estimated using data from the U.S. Environmental Protection Agency’s (EPA) Remote Sensing Information Gateway (RSIG) (US EPA, 2015) (21). Daily averages of PM_2.5_ and ozone were used to calculate monthly averages at the 2010 U.S. Census tract level. Using geocoded residential addresses, PM_2.5_ and O_3_ exposures were linked to each participant's residence and time-weighted based on the duration lived at each address, across two evaluated windows in this study: prenatal and past-year exposures.

**Statistical analyses**

Epigenome-wide association study

Cell-type heterogeneity in nasal tissue was captured using the free-reference ReFACTor algorithm (k=8 based on a scree plot) implemented in the *tca* R package (22). We tested associations between DNAm levels (M-values) at CpGs across the genome with respiratory health outcomes using robust linear regression models implemented in the *limma* R package (23). EWAS were conducted independently for each respiratory outcome, including asthma (ever), current asthma, early-onset asthma, late-onset asthma, total IgE ≥200 IU/ml, FEV_1_ (z-score), FVC (z-score), FEV_1_/FVC, PEF (z-score), and FEF_25-75_ (z-score). Models were adjusted for age, sex, race and ethnicity, body mass index (BMI) (z-score), household income category, smoking and/or vaping in the past month, season of sample collection (sine and cosine), and eight ReFACTor components. Significance was set at a false discovery rate (FDR)<0.05. Probes were annotated to the human GRCh38/hg38 genome assembly using the reannotated Illumina EPICv2 manifest, and those without gene annotation were annotated to the nearest gene using the *rGREAT* R package (24).

Differentially methylated regions (DMRs)

DMRs were estimated using two independent methods implemented in the *ENmix* R Package: *ipDMR* and comb-p (10,25,26). Regions were started at suggestively associated CpGs with FDR<0.1, and those CpGs located within 1000 bp were combined into the same region. All other parameters were set to their default values, and we used as input the summary statistics of the discovery EWAS. We retained DMRs that contain ≥2 CpGs and remain significant after multiple-testing correction at the region level (FDR<0.05). DMRs were annotated to the nearest gene using *rGREAT* (24). To minimize false-positive results, we prioritized DMRs detected using the two software. Genomic regions were considered overlapping across software if they shared ≥50% of their genomic coordinates.

Replication in Project Viva

CpGs and DMRs significantly associated with any respiratory outcome in CHAMACOS at FDR<0.05 were followed up for replication in 540 nasal samples from adolescents (age 13) from the pre-birth cohort Project Viva. Each CpG or DMR was tested only for its association with the same respiratory outcome identified in the discovery analysis. Following the same procedures as in the discovery phase, we tested the association between nasal DNAm (M-values) and respiratory outcomes using *limma* linear robust regression models adjusted for age, sex, race and ethnicity, BMI (z-score), maternal education, smokers at home, season of sample collection (sine and cosine), and cell-type heterogeneity (10 ReFACTor components). For DMRs, we summarized the global DNAm of the region as the mean of M-values of CpGs within it. Mean M-value was used as the outcome of a linear regression model adjusted for the same covariates as in *limma*. The same models were run in CHAMACOS to obtain comparable estimates. Replication was declared for CpGs and DMRs showing the same direction of association as in the discovery phase at *p*<0.05.

DNAm associations with air pollution exposure

CpGs significantly associated with any respiratory outcome were tested for association with air pollution exposures in CHAMACOS. We log_2_-transformed the air pollution exposure variables to minimize the influence of outliers. We tested associations between DNAm at selected CpGs associated with respiratory outcomes and air pollution exposure using linear regression models in R, adjusting for the same covariates as in the main EWAS.

*In-silico* quantitative trait loci analyses

We inspected whether identified CpGs had been associated with nasal epithelial gene expression (expression quantitative trait methylation, eQTM) and/or with genetic variation (methylation quantitative trait loci, meQTL). For that, we queried our CpGs in previous nasal cis-meQTL and cis-eQTM datasets identified in 455 Puerto Rican youth with and without atopic asthma from the EVA-PR study with available whole-genome sequencing (27).

Enrichment analyses

We conducted gene-set enrichment analyses (GSEA) using Enrichr to identify enrichment in biological pathways, gene ontologies, human diseases/traits, transcription factor binding sites, and/or drug signatures (28). We analyzed gene annotations for suggestive signals from any respiratory outcome in the discovery phase using an arbitrary cutoff of FDR<0.2 (943 genes). For each CpG, we prioritized gene annotations based on a re-annotated Illumina EPICv2 manifest based on GENCODEv47. For those without gene annotations, we used the nearest single gene annotation from *rGREAT* (24). For CpGs annotated to multiple genes, we prioritized the annotation that overlaps the two methods. We queried the databases GWAS Catalog 2025, MSigDB Hallmark 2020, KEGG 2026, ChEA 2022, and DSigDB. To ensure the robustness of associations, we prioritized enrichment terms that remained significant (*p*<0.05) after reassessing GSEA by varying alternative gene-selection cutoffs (FDR<0.1, 214 genes; and FDR<0.3, 3,058 genes).

Epigenetic aging

We used the methscore() *ENmix* function to calculate the first-generation Horvath and Skin&Blood clocks (29), given that they were trained in multiple tissues, including epithelial cells. We tested associations between epigenetic age and respiratory outcomes using linear regression models adjusted for age, sex, race and ethnicity, BMI (z-score), household income category, smoking and/or vaping in the past month, and season of sample collection (sine and cosine). Significance was declared for each outcome at *p*<0.025 (α=0.05/2 epigenetic clocks=0.025). We conducted a sensitivity analysis adjusted for ReFACTor components.

**SUPPLEMENTARY FIGURES**

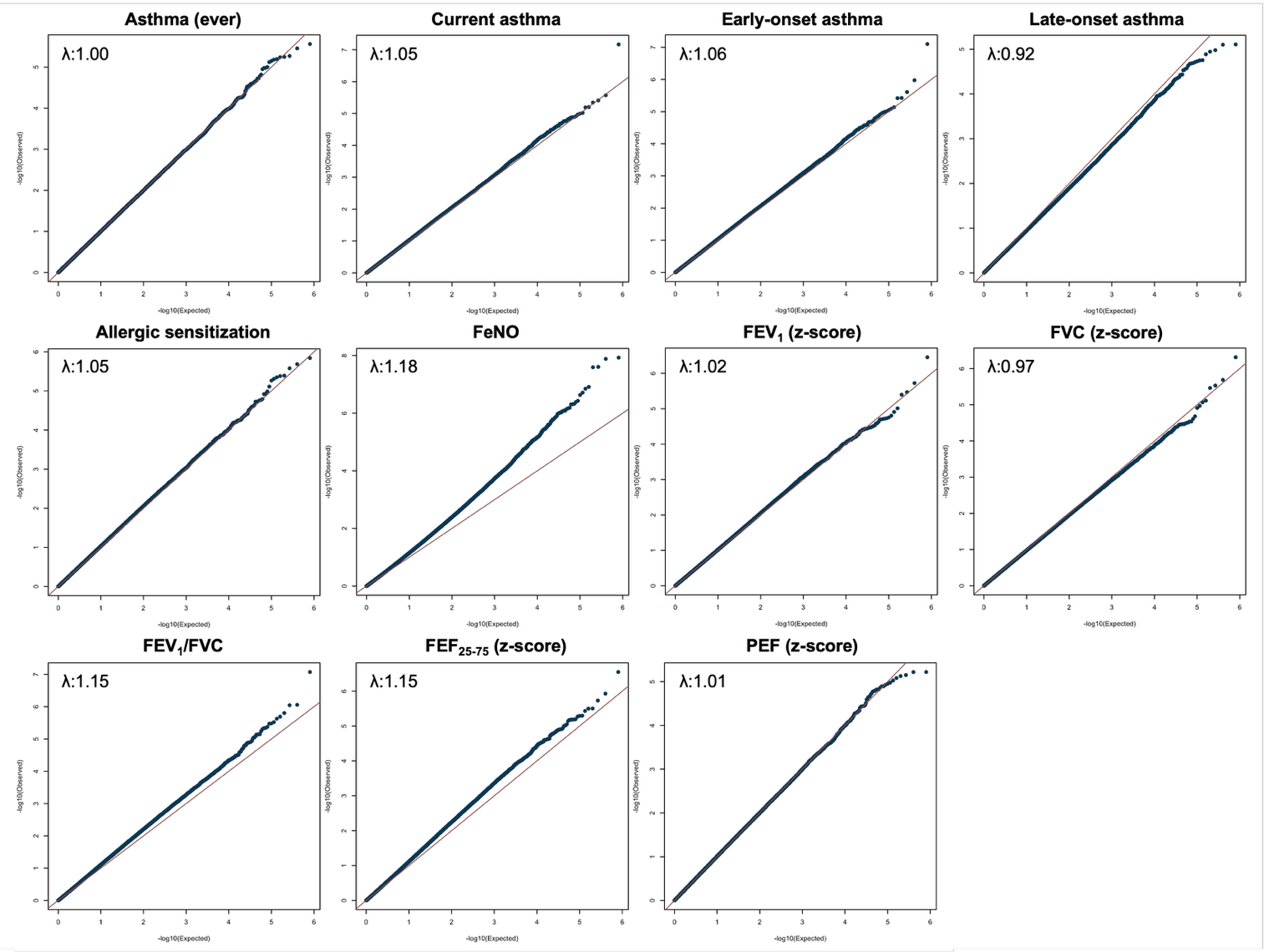

**Supplementary Figure S1.** QQ-plots of the EWAS for the 11 respiratory outcomes analyzed in this study.

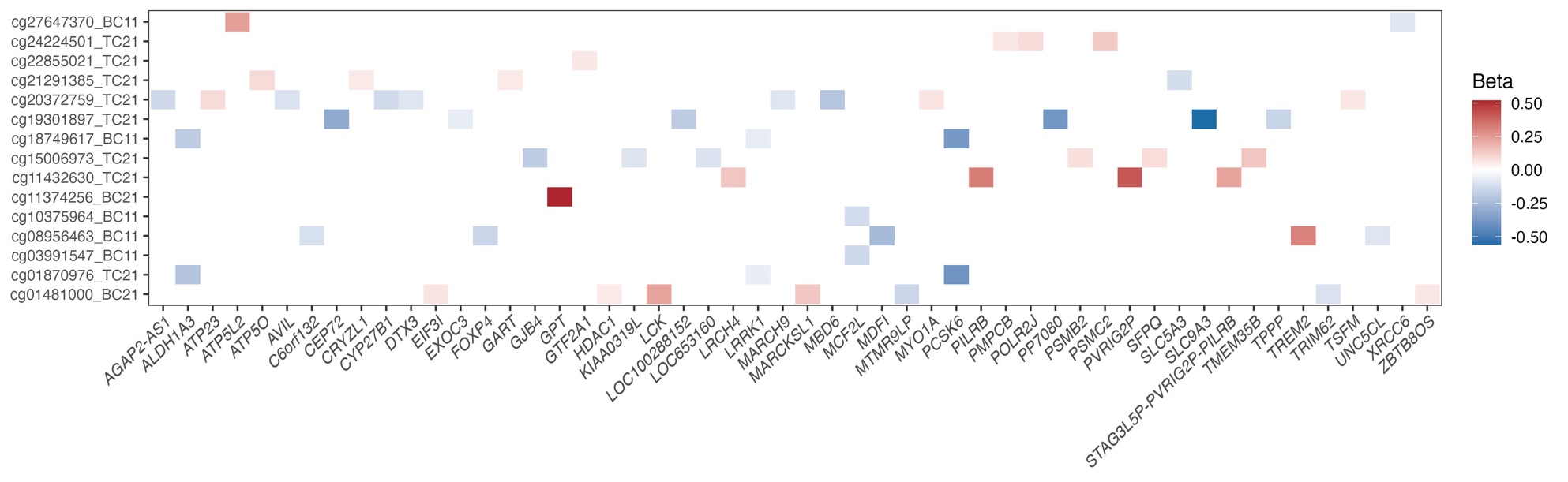

**Supplementary Figure S2.** Heatmap summarizing the in-silico expression quantitative trait methylation (eQTM) analyses based on a public database from EVA-PR (Epigenetic Variation and Asthma in Puerto Ricans) cohort using nasal epithelial DNA methylation and gene expression data. CpGs significantly associated with FeNO in our study are represented on the y-axis, and genes whose nasal epithelial gene expression levels are in silico associated with these CpGs are shown on the x-axis. Up-regulation is shown in red, and down-regulation in blue. Only significant associations (FDR<0.05) are colored (multiple comparisons were adjusted in the original database considering comparisons across the whole genome).

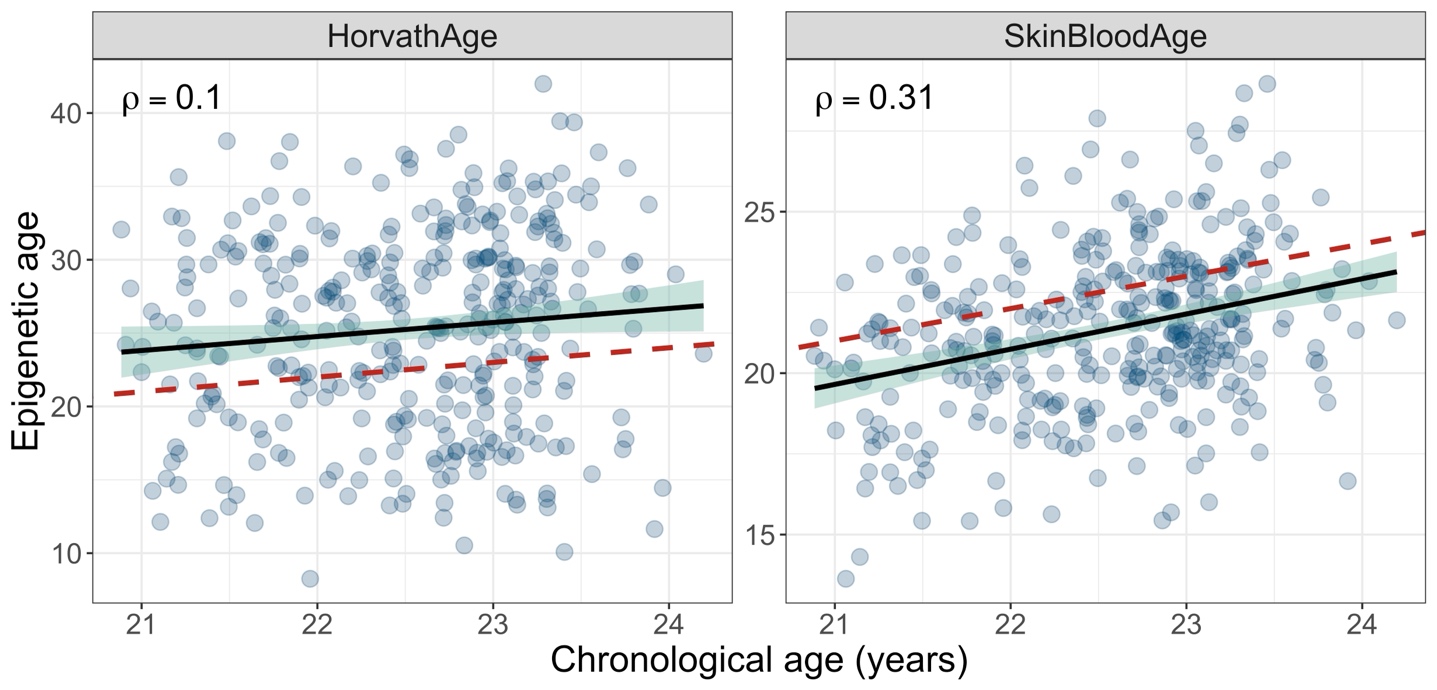

**Supplementary Figure S3.** Scatter plot showing the correlation between chronological age (x-axis) and epigenetic age (y-axis). Epigenetic age was calculated from nasal DNA methylation data using the HorvathAge (left panel) and Skin&BloodAge (right panel) epigenetic clocks. The black line represents the slope and the 95% confidence interval (green shadow). The dashed red line represents the 1:1 relationship. The correlation between chronological and epigenetic age was assessed using Pearson’s correlation coefficient
